# A Predictive Internet-Based Model for COVID-19 Hospitalization Census

**DOI:** 10.1101/2020.11.15.20231845

**Authors:** Philip Turk, Thao Tran, Geoff Rose, Andy McWilliams

## Abstract

The COVID-19 pandemic has strained hospital resources and necessitated the need for predictive models to forecast patient care demands in order to allow for adequate staffing and resource allocation. Recently, other studies have looked at associations between Google Trends data and the number of COVID-19 cases. Expanding on this approach, we propose a vector error correction model (VECM) for the number of COVID-19 patients in a healthcare system (Census) that incorporates Google search term activity and healthcare chatbot scores. The VECM provided a good fit to Census and very good forecasting performance as assessed by hypothesis tests and mean absolute percentage prediction error. Although our study and model have limitations, we have conducted a broad and insightful search for candidate Internet variables and employed rigorous statistical methods. We have demonstrated the VECM can potentially be a valuable component to a COVID-19 surveillance program in a healthcare system.

## Introduction

The SARS-CoV-2 coronavirus, initially emerging in Wuhan, China on December 2019, has spread worldwide in what is now described as the COVID-19 pandemic. The coronavirus outbreak was declared a public health emergency^1^ by the World Health Organization [WHO] in early 2020, and as of October 17, there were over 39 million confirmed cases worldwide with over a million lives lost^2^. International guidelines and regional restrictions in activities have been put in place to slow the spread of the virus^3,4^. While evidence supports the effectiveness of these actions in containing the spread of SARS-CoV-2 (“flattening the curve”), the health and economic consequences have been devastating on many levels^5,6^.

At the conclusion of summer 2020, the US has had more COVID-19 cases and deaths than any other country^2^. As of June 30, the US has 4% of the world’s population, but 25% of its coronavirus cases. While most states avoided a rapid surge in cases during the first phase of the pandemic, the majority of them have begun to lift social distancing and gathering restrictions, raising concern that we may see increased infection incidence and mortality rates^7–9^. Indeed, after relaxing measures, several states (e.g., California, Texas, Arizona, Florida) saw very large surges in the number of new COVID-19 cases^2^. Without a widely available vaccine, we expect that the pandemic activity will continue to rise and fall through the winter, requiring health care systems to remain vigilant as they balance hospital resources.

As has been seen in this pandemic, when SARS-CoV-2 prevalence grows quickly and reaches high levels in a community, large numbers of people develop symptomatic COVID-19 infection. Many require hospitalization, and this has the capacity to overwhelm regional health care resources (e.g., Northern Italy and NY). Acknowledging this risk, health care systems have implemented crisis planning to guide infection management, bed capacity, and secure vital supplies (e.g., ventilators and personal protective equipment)^10–12^. Ideally, health system’s efforts to best prepare for COVID-19 demand surges would be informed by data that provide early warning, or “lead time”, on the local prevalence and impact of COVID-19.

As the pandemic has progressed, traditional epidemiological models (e.g., Susceptible-Infected-Removed model) have not consistently provided health system leaders with this lead time. In searching for other leading indicators, researchers have turned to internet data that reflect trends in community behaviors and activity. In recent years, researchers have utilized various internet search data to predict different health-related topics, such as dengue incidence^13^, infectious disease risk communication^14^, influenza epidemic monitoring^15^, and malaria surveillance^16^. Google Trends is one of the most popular tools that allows researchers to pull search query data of a random, representative sample drawn from billions of daily searches on Google and Google-associated search engines^17^. Rather than a raw count of “hits”, Google Trends data reflects the relative popularity of a search term, or relative search volume (RSV), over a specified time range and location.

In the last six months, several papers have made use of Google Trends data to test the association between the RSV of certain coronavirus-related terms and the number of cases and deaths related to COVID-19^18–21^. While there are methodological concerns that need to be considered when using Google Trends RSVs to build a predictive model for COVID-19 modeling applications^20,22^, these papers have contributed to our collective understanding of the relationship between the public’s internet search behavior and the pandemic, supporting the notion that search query data can be used for surveillance purposes.

In addition to Internet users’ search query data, another source of data that is of importance for public health research is geospatial health data. Since the initial outbreak of COVID-19 in Wuhan, China, researchers have believed that population mobility is a major driver of the exponential growth in the number of infected cases^23–25^. It is now well-accepted that mobility reduction and social distancing are timely and effective measures to attenuate the transmission of COVID-19^26,27^. Thus, it stands to reason that mobility changes may be a predictor of COVID-19 case volume. Many different interactive dashboards are available and display up-to-date regional mobility data that are publicly available, most notably from Facebook and Apple Maps^28–32^. In this paper, we specifically considered Apple Mobility Trend Reports and Facebook Movement Range Maps, which are mobility data reported in the form of aggregated, privacy-protected information, to look for potential “hot spots” or trends in infection as shelter-in-place orders are lifted and more people travel and mix together.

Another area with great potential in modeling COVID-19 cases is the data generated by virtual AI-based triage systems (also known as “healthcare chatbots”). During the COVID pandemic, these chatbots have been deployed to provide user-friendly, conversational consultation to people who are concerned they may have SARS-CoV-2^33,34^. In particular, Microsoft offers its Health Bot service to healthcare organizations^35^. Medical content, together with an interactive symptom checker, custom conversational flow, and a system of digital personal assistants can be integrated into the Health Bot configuration to help screen people for potential coronavirus infection through a risk assessment and determine an appropriate course of action depending on the result^36–39^.

As people take the online COVID-19 risk assessment within a healthcare system, Health Bot user outcomes (with no personally identifiable information) can be compiled. The number of people “flagged” with COVID-19 can then be potentially used to predict new cases of COVID-19 in the near future. Specifically, if a hospital has its own Health Bot for delivering a tele-health COVID-19 risk assessment to the public, then it is reasonable to expect that people who are identified as having COVID-19 are likely to seek treatment from the same hospital.

Atrium Health is a healthcare system operating across North Carolina, South Carolina, and Georgia, with the majority of its hospitals located in the greater Charlotte metropolitan area. Investigators from the Atrium Health Center for Outcomes Research and Evaluation sought to leverage data from internet search trends, mobility, and Health Bot interactions to provide leadership with information that would allow for planning purposes during the pandemic. In considering what candidate “Internet variables” to use, we adopted several inclusion criteria.

First, the data must be free. Second, the data must be current, so that decisions can be made in near real-time. Lastly, we also required that the data are publicly available and easy to access.

This paper describes the steps to characterize and understand the relationships between our Internet variables and the daily total number of COVID-19 patients hospitalized in our hospital system’s primary market. Furthermore, we sought to develop a forecast model for these patients to provide advance warning of any anticipated surges in patient care demands.

## Methods

### Measures

Our interest lies in the population served by Atrium Health’s greater Charlotte market area which spans approximately 11 counties in western North Carolina and two counties in northern South Carolina. This area includes approximately 400,000 South Carolina residents, 2.5 million North Carolina residents (24% of the North Carolina population), over 1.1 million of which live in Mecklenburg County and 900,000 within North Carolina’s largest city, Charlotte^40^.

Because of the focus on health care system capacity, our outcome variable of interest is the total COVID-19 positive census across 11 Atrium Health Hospitals that serve the greater Charlotte market (hereafter referred to as “Census”) with an additional virtual hospital, Atrium Health Hospital at Home, providing hospital level care in a patients home. Census is a cross-sectional count taken each morning as the total number of patients hospitalized and COVID-19 positive.

Google Trends data represents the relative search popularity of different topics. Specifically, the RSV of a search term is calculated as the proportion of interest in that particular topic relative to all searches over a specified time range and location. The RSV is normalized to a scale of 0 to 100. “0” indicates that the term appears in very few searches and “100” shows maximum interest in the term for the chosen time range and region^17^. To retrieve Google Trends data for our analysis, we utilized the gtrendsR package in R (https://cran.r-project.org/web/packages/gtrendsR/gtrendsR.pdf). We performed twelve different queries from 02/21/20 to 08/01/20 for Google Trends’ “Charlotte NC” metro designation (county-level data is unavailable) using a list of terms obtained based on our prior beliefs and expertise. Since punctuations can influence the search results, we followed the guidelines from Google News Initiative^41^ to refine our search queries. Details on the search terms can be found in Table 1.

**Table 1:** Google Trends Search Terms Used

| covid | coronavirus |
| --- | --- |
| symptoms covid | covid testing + covid test + covid19 testing + covid19 test + covid 19 testing + covid 19 test |
| fever + chills | headache |
| “shortness of breath” | “shortness of breath” + “trouble breathing” + “difficulty breathing” |
| tired + fatigue | pneumonia |
| “sore throat” + cough | CDC |

*Google Trends search terms with no punctuation will contain the term(s), along with other words, in any order. Using double quotation marks give results that include that exact term, possibly with words before and after. The use of a plus sign (+) is shorthand for OR.*

Apple Mobility Trend Reports collect Apple Maps direction requests from users’ devices and record the relative percentage change in driving direction requests compared to the baseline requests volume on January 13, 2020 on a daily basis. These data are available at the county-level allowing us to pull data specifically for Mecklenburg County, North Carolina from 02/21/20 to 08/01/20. For unknown reasons, data were missing for two days (May 11 and May 12), so we replaced them with estimates using linear interpolation.

Facebook Movement Range Maps include data from Facebook users who access Facebook on a mobile device, with Location History and background location collection enabled. A data point for a given region is computed using the aggregate locations of users for a particular day. Specifically, there are two metrics, Change in Movement and Staying Put, that provide slightly different perspectives on movement trends. The Change in Movement metric measures the proportion change in frequency of travel (relative to the day of the week) compared to the last two weeks of February recorded on a daily basis, while the Staying Put metric measures the proportion of the regional population who remained in one location for 24 hours. Once again, we pulled data from 02/21/20 to 08/01/20 for Mecklenburg County, North Carolina.

In the early days of the pandemic, Atrium Health collaborated with Microsoft Azure to launch its own public-facing Health Bot to converse with people about their COVID-19 symptomology. Generally, a person will respond “Yes/No” to a series of questions on COVID-19 symptoms, whether they belong to a vulnerable group (e.g., elderly people, pregnant women, people with compromised immune system, etc.) and whether they are scheduled for a medical procedure or surgery. Depending on the users’ answers, the Health Bot will use branched logic to indicate if the person is at risk of having COVID-19 and prompt appropriate further actions. In this study, we focused on the number of times that people are flagged as “may have COVID-19” for further analysis. These data are daily counts of users that have completed the risk assessment and that Health Bot has classified as “may have COVID-19”.

After the data were pulled, we generated 16 time plots (12 for Google Trends, 1 for Apple, 2 for Facebook, and 1 for Health Bot). We then computed Spearman’s correlation coefficient for Census at time *t* and each of the “lagged” Internet variables at times *t, t* – 1,…, *t* – 14. A lag of -14 was chosen because 14 days is consistent with the known maximum incubation period associated with COVID-19^42^. For each variable, we looked for the maximum absolute correlation coefficient across all 15 values to guide the selection of the most important variables for further study.

### Analytic Approach

In considering models for observed time series, suppose we have the stochastic process {*y*_*t*_: *t* = 0, ±1, ±2, …}. Note that *y*_*t*_ is oftentimes referred to as the “level of the time series”. A stochastic process {*y*_*t*_} is (weakly) stationary if the mean *E*[*y*_*t*_] is constant over time, and if the autocovariance *Cov*[*y*_*s*_, *y*_*t*_] = *Cov*[*y*_*s+k*_, *y*_*t+k*_] for all times *s* and *t*, and lags *K* = 0, ±1, ±2, …. Informally, a stationary time series is one whose properties do not depend on the time at which the series is observed. Thus, time series with non-constant trends, seasonality, changes in variance, etc., are nonstationary. We used the methodology described in Pfaff^43^ and Dickey and Fuller^44^ to determine whether or not a time series is stationary. If it is not, then we further characterize the nature of the nonstationarity.

Suppose *y*_*t*_ can be decomposed into a deterministic linear trend component and a stochastic residual component that is an autoregressive-moving average (ARMA) process. A time series can exhibit a type of nonstationarity, perhaps confusingly, referred to as “difference-stationary”, which means that *y*_*t*_ **−** *y*_*t-*1_ is a stationary stochastic process. Also, a time series can exhibit a type of nonstationarity referred to as “trend-stationary”. Once the data are detrended, the resulting residual time series is a stationary stochastic process. The difference between these two types of nonstationarity may imply different time series dynamics and hence, different forecasts.

In order to understand the model proposed in this research, we must first define cointegration. We use a broader definition^45^ than is typically defined elsewhere in the literature. Specifically, let ***y***_*t*_ be an *n* × 1 vector of variables ***y***_*t*_. Note that ***y***_*t*_ can contain time series that are either difference-stationary *or* trend-stationary. This vector is said to be cointegrated if there exists an *n* × 1 vector ***β***_*i*_ (**≠ 0**) such that ***β*′**_*i*_***Y***_***t***_ is trend-stationary. ***β***_*i*_ is then called a cointegrating vector. In fact, it is possible that there are *r* linearly independent vectors ***β***_*i*_ (*i* = 1, …, *r*).

We now consider some background behind our model. A vector autoregression model of order *K* (VAR(*K*)) is defined as:

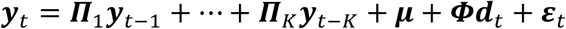

where *t* = 1, …, *T*. Here, ***y***_*t*_ is an *n* × 1 vector of time series at time *t*, ***П***_*i*_ (*i* = 1, …, *k*) is an *n* × *n* matrix of coefficients for the lagged time series, ***μ*** is an *n* × 1 vector of constants, ***d***_*t*_ is an *p* × 1 vector of deterministic variables (e.g., seasonal indicators, time, etc.), and ***Φ*** is a corresponding *n* × *p* matrix of coefficients. We assume the *ε_t_* are independent *n* × 1 multivariate normal errors with mean **0** and covariance matrix **Σ**. In order to determine a value for *K* in practice, one can sequentially fit a VAR model, for *k* = 1, …, 10, say, and compare Akaike’s Information Criterion (AIC) values^46^, where smaller values of AIC offer more evidence to support a specific model^47^.

One way to respecify the VAR model is as a (transitory) vector error correction model (VECM). Using linear algebra, we can obtain:

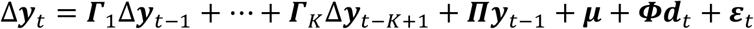

where *Δ****y***_*t*_ is the (first) difference ***y***_*t*_ **−** *y*_*t-*1_, *****Γ*****_*i*_ = **−**(***П***_*i*+1_ *+* ⋯ *+* ***П***_*k*_), for *i* = 1, …, *k* **−** 1 and *k* ≥ 2, and ***П*** = **−**(*I* **− *П***_1_ **− ⋯ − *П***_*k*_) for an identity matrix ***I*** of order *n*. In effect, a VECM is a VAR model (in the differences of the data) allowing for cointegration (in the levels of the data). The matrix ***П*** measures the long-run relationships among the elements of ***y***_*t*_, while the ***Γ***_*i*_ measure short-run effects. ***Пy***_*t-*1_ is oftentimes called the “error correction term” and it is assumed this term is stationary. More rigorous background on cointegration, the VAR model, and the VECM can be found in Pfaff^43^.

An important part of fitting a VECM is determining the number (*r*) of cointegrating relationships that are present. It can be shown that the rank of the matrix ***П*** is equal to *r*. In practice, the most interesting case is when *r* ∈ (0, n). In this case, we can use a rank factorization to write, ***П*** = ***αβ*′**, where both ***α*** and ***β*** are of size *n* × *r*. Therefore, ***Пy***_*t-*1_ = ***αβ*′***y*_*t-*1_ is stationary. Because ***α*** is a scale transformation, ***β*′***y*_*t-*1_ is stationary. By our definition of cointegration, there are *r* linearly independent columns of ***β*** that are the set of cointegrating vectors, with each of these column vectors describing a long-run relationship among the individual time series. Elements in the vector ***α*** are often interpreted as “speed of adjustment coefficients” that modify the cointegrating relationships. The number of cointegrating relationships can be formally determined using Johansen’s procedure^48^.

Following Johansen^49^ and Johansen and Juselius^50^, we consider how to specify the deterministic terms in the VECM using AIC and a likelihood ratio test on linear trend. In our case, due to the nature of the research problem and by visual inspection of the time plots, we initially set ***Фd***_*t*_ = **0** (this form of the model is known as a restricted VECM). We then consider two possibilities for the constant ***μ***. The first possibility is to place ***μ*** inside the error correction term. Specifically, define an additional restriction ***μ*** = ***αρ***. Then, the error correction term can be rewritten as ***α***(***β*′***y*_*t-*1_ *+* ***ρ***) so that the cointegrating relationships have means, or intercepts, ***ρ***. The second possibility is to leave ***μ*** as is to account for any linear trends in the data.

We used maximum likelihood estimation to fit the VECM and report estimates and standard errors for elements of ***α, β*** and ***Γ***_1_, along with corresponding *t*-tests run at a significance level of 0.05.

When fitting a VECM, it is important to check the goodness-of-fit. Using the fitted VECM, and *r*, as determined by Johansen’s procedure, we backed out estimates of the coefficients ***П***_*i*_ of the corresponding VAR model of order *K* (in levels). This was then recast as a VAR model of order 1; that is, it was rewritten in “companion matrix” form^51^. The VECM is stable, i.e., correctly specified with stationary cointegrating relationships, if the modulus of each eigenvalue of the companion matrix is strictly less than 1. Another stability check is to investigate the cointegration relationships for stationarity. For the later, we again used the methodology described in Pfaff^43^ and Dickey and Fuller^44^.

Residuals diagnostics were run to check assumptions on the errors ε_*t*_. We computed a multivariate Portmanteau test for serially correlation, and generated autocorrelation function (acf) and cross-correlation function (ccf) plots to guide interpretation. Also, we computed univariate and multivariate Jarque-Bera tests for normality.

For a VECM, predictions and forecasts for the level of a time series are obtained by transforming the fitted VECM to its VAR form. It can be shown that in-sample (training) predictions are actually one-day-ahead forecasts using estimated model coefficients based on the whole time series. We obtain approximate in-sample prediction intervals by making use of the estimated standard deviation of the errors taken from the Census component of the model. Out-of-sample (test) forecasts are computed recursively using all three time series from the VAR model fit to past data, for horizons equal to 1, 2,…, 7, say. The construction of out-of-sample forecast intervals as a function of the horizon are described elsewhere in the literature^52^.

In order to assess the out-of-sample forecasting performance of our VECM, we used a time series cross-validation procedure. In this procedure, there is a series of test sets, each consisting of 7 Census observations. The corresponding training set consists only of observations that occurred prior to the first observation that forms the test set. Thus, no future observations can be used in constructing the forecast. We gave ourselves a 2-week head start on the frontend of the Census time series, and a 1-week runway on the backend. For the 88 days starting from 04/29/20 up to 07/25/20 by one day increments, we iteratively fit the VECM and computed the 7-days-ahead out-of-sample mean absolute percentage prediction error (MAPE). MAPE is defined here as 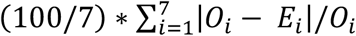, where *O*_*i*_ is the observed Census value, *E*_*i*_ is the projected Census value, and *i* = 1, 2, …, 7 horizons. Notice the “origin” at which the forecast is based, and which delineates training versus test set, rolls forward in time. We chose 7 days because it is in accordance with the weekly cadence of reporting on pandemic behavior and forecast metrics at Atrium Health. In addition, 7 days is a reasonable average timeframe for infection with coronavirus, incubation, and the potential subsequent need for hospitalization. As a baseline for comparison, we also evaluated our VECM against a basic ARIMA model, derived using the approach of Hyndman and Khandakar^53^, using the same time series cross-validation procedure.

All data analysis was done using R statistical software, version 3.6.2, with the packages `tsDyn`, `vars`, and `urca` being the more important ones for fitting the VECM. The data and code used in the data analysis is publicly available at GitHub (https://github.com/philturk/CovCenVECM).

## Results

The 16 time plots for the Internet variables are shown in Figure 1. The first three rows are for those from Google Trends, while the last row contain those from Apple, Facebook, and Health Bot. Clearly, several of the time series are visibly nonstationary.

**Figure 1:**
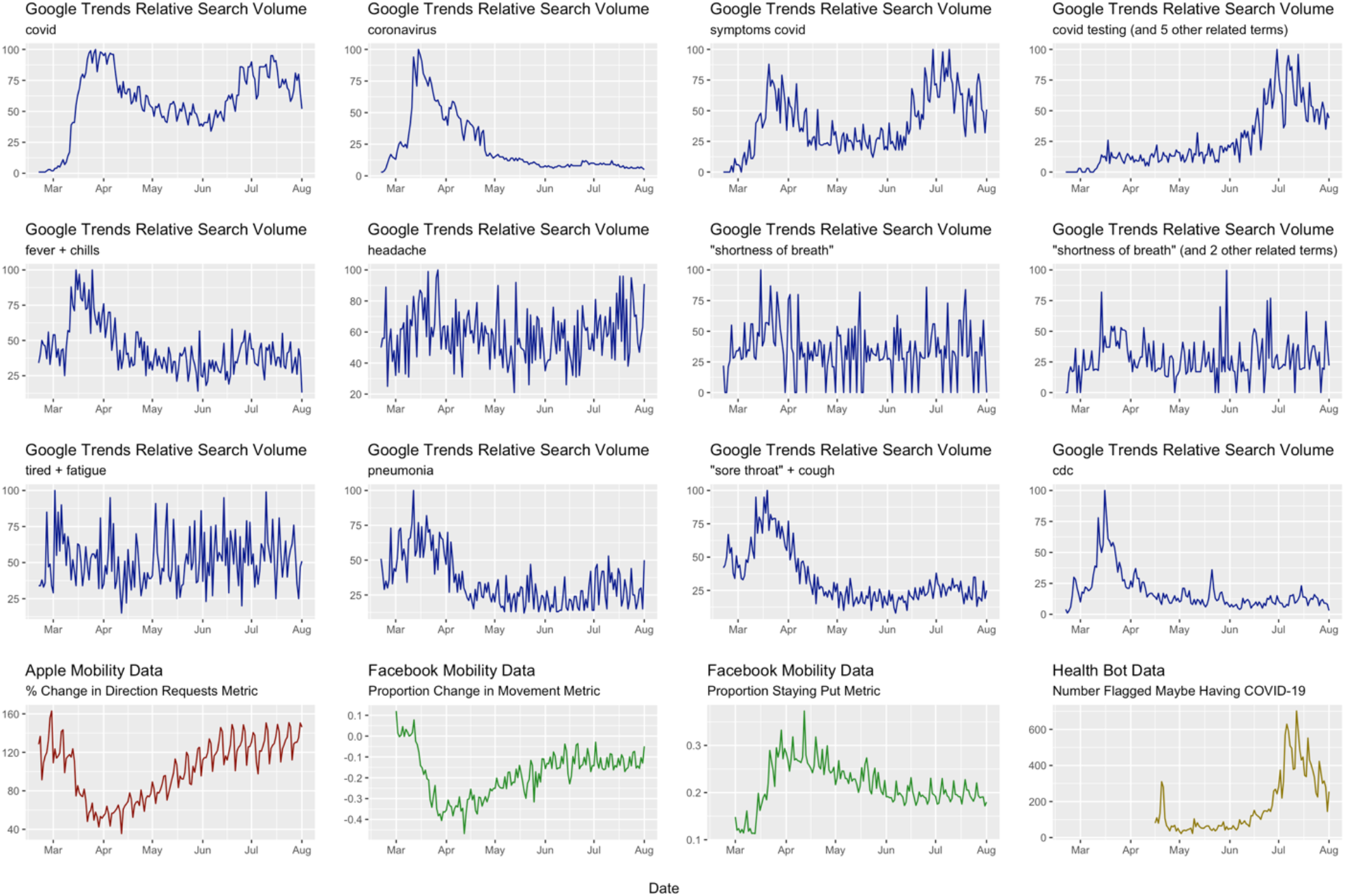
Time Plots for Internet Variables Used in Data Analysis

In looking at the maximum absolute Spearman’s correlation coefficient between Census and each Internet variable across lags 0, -1,…, -14, two variables stood out (Table 2). The first was Health Bot, with a maximum absolute correlation coefficient of 0.865 at time *t* – 4. The second was the Google Trends search term for covid testing + covid test + covid19 testing + covid19 test + covid 19 testing + covid 19 test, henceforth, referred to as Testing. Testing had a maximum absolute correlation coefficient of 0.819 at time *t*. Three other search terms were noted, but their maximum absolute correlation coefficients were substantially lower than HealthBot and Testing. For covid and symptoms covid, it was felt their searches might substantially overlap with Testing. The search term coronavirus had a negative correlation, likely attributable to people’s initial interest in the novelty of COVID-19, which waned over time, as reflected from the beginning of June onward when RSV values for coronavirus were quite small. Therefore, for the sake of parsimony, the three other search terms were not considered further for this research.

**Table 2:** Internet Variables and Maximum Absolute Correlation with Census in Previous 14 Days.

| Internet Variable | Lag<br>(in days from time $t$) | Maximum Correlation<br>(in absolute magnitude) |
| --- | --- | --- |
| covid | -13 | 0.518 |
| coronavirus | 0 | -0.565 |
| symptoms covid | -14 | 0.614 |
| covid testing<br>(and 5 other related terms) | 0 | 0.819 |
| fever + chills | 0 | -0.322 |
| headache | -5 | 0.083 |
| “shortness of breath” | 0 | -0.073 |
| “shortness of breath”<br>(and 2 other related terms) | -13 | 0.156 |
| tired + fatigue | -13 | 0.125 |
| pneumonia | -1 | -0.453 |
| “sore throat” + cough | 0 | -0.478 |
| cdc | 0 | -0.448 |
| Apple mobility | 0 | 0.378 |
| Facebook movement | -14 | 0.153 |
| Facebook staying put | -14 | -0.109 |
| Health Bot | -4 | 0.865 |

After examination of scatter plots, and in preparation for modeling, we transformed both Health Bot (by taking the natural logarithm) and Testing (by taking the square root) to linearize the relationship between each of these variables and Census. We then generated longitudinal “cross-correlation”-type of profiles for Health Bot and Testing using Pearson’s correlation coefficients for lags 0, -1,…, -14 as shown in Figure 2. We can see strong correlations, all well above 0.80, throughout the time period under consideration.

**Figure 2:**
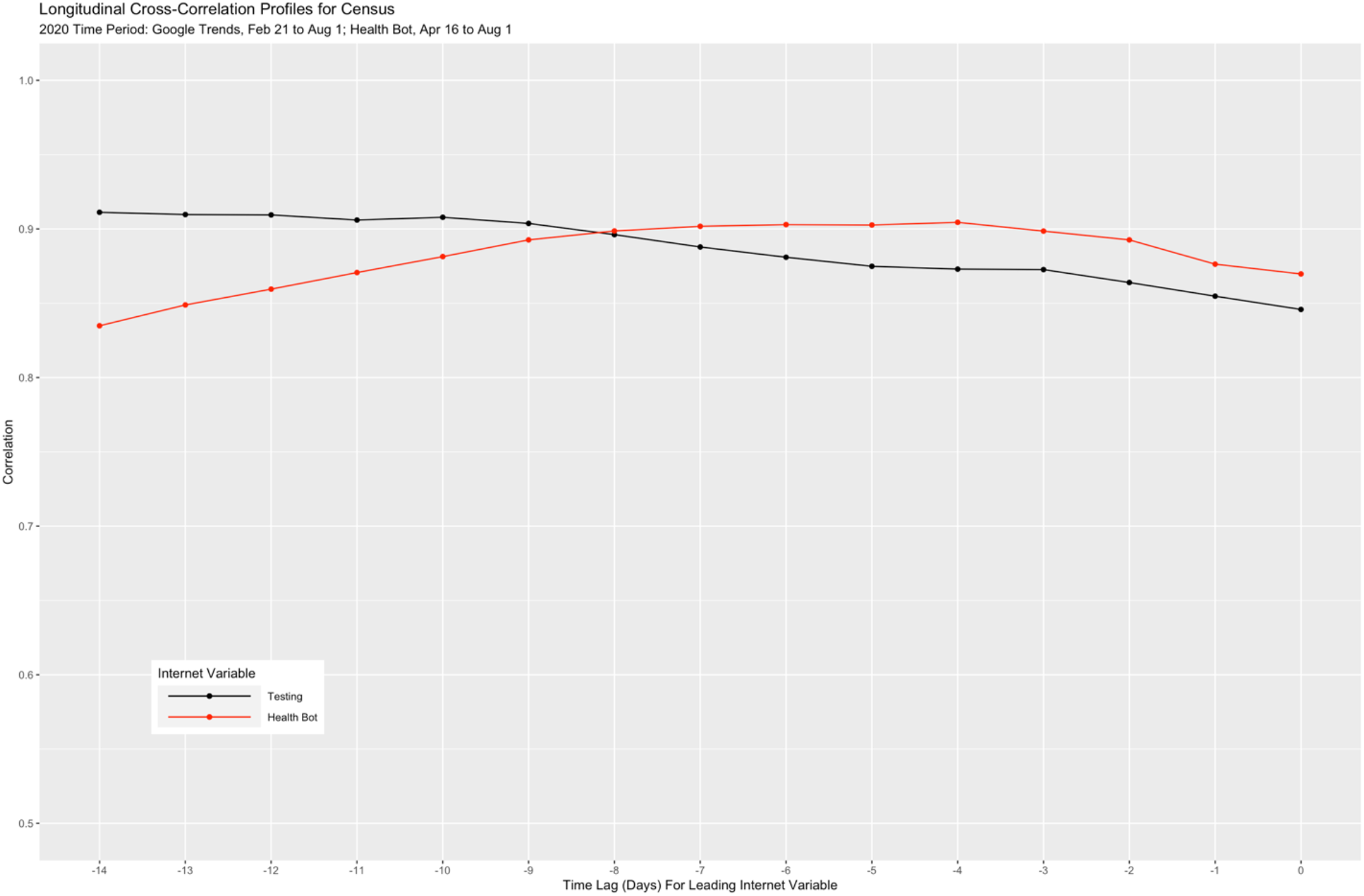
Longitudinal Cross-Correlation Profiles for Census and the Two Internet Variables Health Bot and Testing

To better understand the relationships among the three time series, we normalized both the Health Bot and Census time series to the same [0, 100] scale as Testing, and obtained the results in Figure 3. Both the Testing and Health Bot time series appear to share common features of the Census time series (e.g., approximate linear increase from mid-May until mid-July). There is also the suggestion that both the Testing and Health Bot time series “lead” the Census time. For example, from mid-April until the beginning of May, Health Bot shows a downward linear trend and this behavior is mirrored in the Census time series roughly one week later.

**Figure 3:**
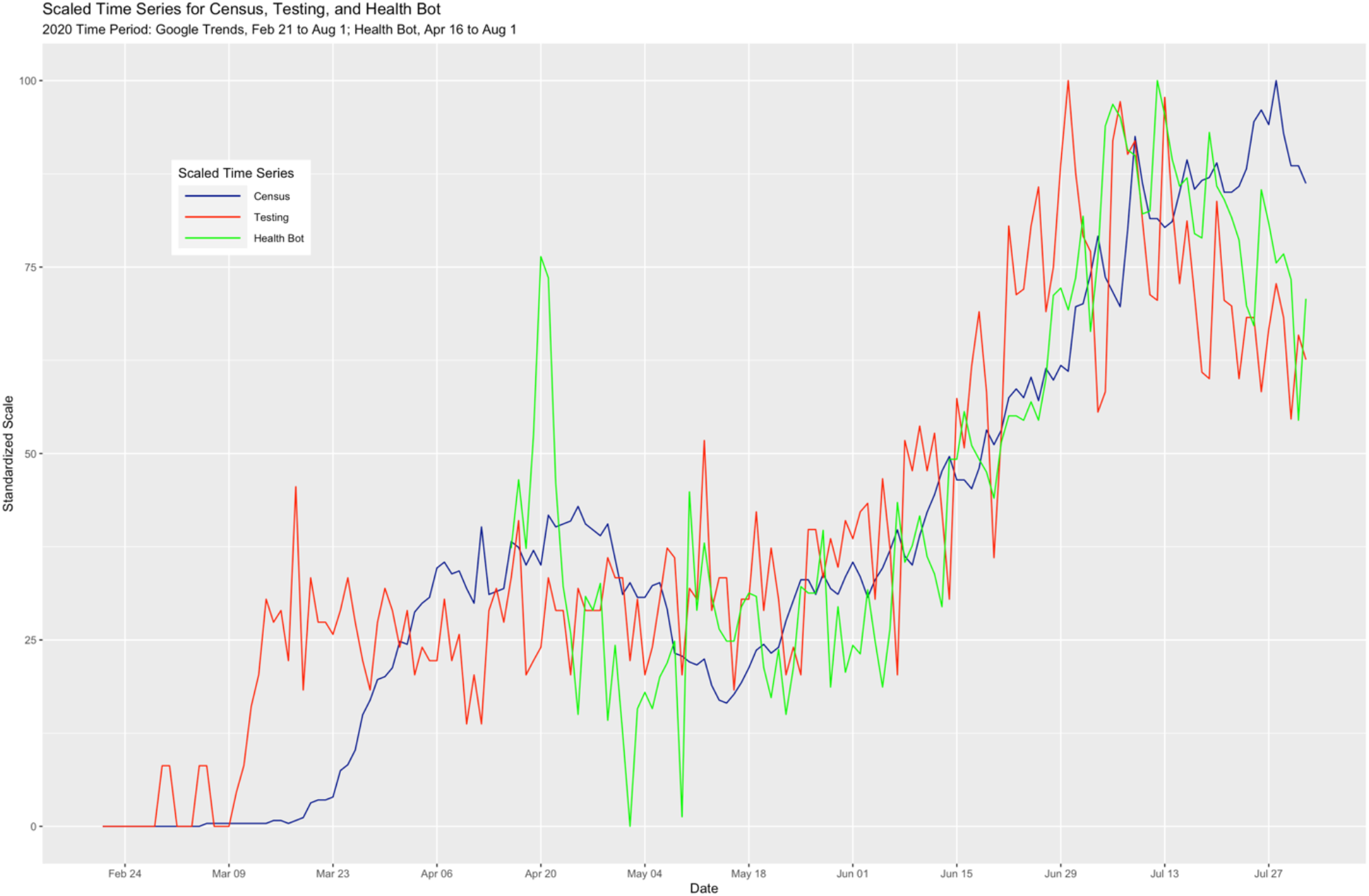
Multivariate Time Series Plot for Census and the Two Internet Variables Health Bot and Testing

Using the methodology of Pfaff^43^ and Dickey and Fuller^44^, all three time series are nonstationary. Specifically, the Census time series is difference-stationary, while the Health Bot and Testing time series are both trend-stationary.

Results from examining AIC values after fitting a VAR model to Census, Testing, and Health Bot, sequentially increasing the lag order up to 10, were inconclusive. Therefore, we chose the minimum value of *K* = 2. Johansen’s procedure (using the trace test version) indicated that two cointegrating vectors should be used. A comparison of the two AIC values for restricted VECM models described in the Methods suggested placing ***μ*** inside the error correction term (AIC = 212.598) as opposed to not doing so (AIC = 217.592). This was also corroborated by a likelihood ratio test for no linear trend (*p*-value = 0.32).

For the sake of brevity, and because we are most interested in modeling Census, we only show the portion of the fitted VECM pertaining to Census. Both ***α*** and ***β*** are not unique, so it is typical in practice to normalize them. The normalization we used is the Phillips triangular representation, as suggested by Johansen^49^. The expression for Census in scalar form using general notation is:

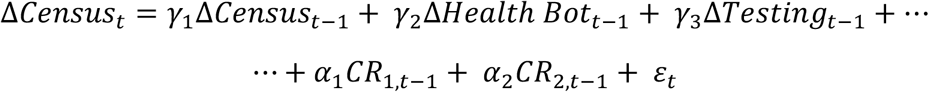

where *γ*_1_, *γ*_2,_ and *γ*_3_ are the corresponding elements of ***Γ***_1_, ***α***_1_ and ***α***_2_ are the corresponding elements of *α*, and *CR*_1_ and *CR*_2_ are the first and second cointegrated relationships. Collectively, ***α***_1_*CR*_1,*t-*1_ and ***α***_2_*CR*_2,*t-*1_ are the error correction terms. In our case, we obtained the results shown in Table 3:

**Table 3:** Results from Fitted VECM.

| Parameter | Estimate | Standard Error |
| --- | --- | --- |
| $\gamma_1$ | 0.0424 | 0.0931 |
| $\gamma_2$ | -3.8514 | 2.5773 |
| $\gamma_3$ | -1.3412 | 0.8382 |
| $\alpha_1$ | 1.6559* | 0.7837 |
| $\alpha_2$ | 4.3317* | 2.1967 |

*Results denoted with an asterisk are statistically significant at a significance level of 0.05. The estimates for $\alpha_1$ and $\alpha_2$ are normalized.*

An overall omnibus test for the Census component of the VECM was statistically significant (*F*_0_ = 3.393 on 5 and 101 degrees of freedom; *p*-value = 0.0071). We see that the long-run effects for both cointegrated relationships were important in modeling the first difference of Census at time *t*. However, the short-run, transitory effects as measured by first differences of Census, Health Bot, and Testing at lag 1 were not statistically significant.

Furthermore, the expressions for the cointegrated relationships are:

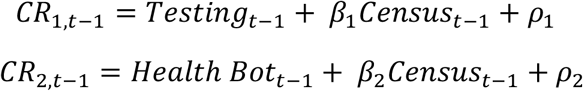

where *ρ*_1_ and ***ρ***_2_ are the corresponding elements of ***ρ***, and *β*_1_ and *β*_*2*_ are the corresponding elements of ***β***. We obtained the results shown in Table 4:

**Table 4:** Results from Fitted VECM.

| Parameter | Estimate |
| --- | --- |
| $\rho_1$ | -1.9911 |
| $\rho_2$ | -2.9994 |
| $\beta_1$ | -0.0257 |
| $\beta_2$ | -0.0131 |

*The estimates for all parameters are normalized.*

Considering the model, parameter estimates from the previous two tables, and looking at *CR*_1,*t-*1_, we see that if Testing is unusually low relative to Census at time *t* **−** 1, so that *Testing*_*t-*1_ *<* **−**0.0257*Census*_*t-*1_ **−** 1.9911, then this suggests a decrease in Census at time *t*. Similarly for *CR*_2,*t-*1_, if Health Bot is unusually low relative to Census at time *t* **−** 1, so that *Health Bot*_*t-*1_ *<* **−**0.0131*Census*_*t-*1_ **−** 2.9994, then this suggests a decrease in Census at time *t*.

A check of the modulus of all the eigenvalues from the companion matrix associated with the VECM showed them all to be well below 1, suggesting stability of the model. Inspection of the two fitted cointegration relationships 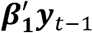 and 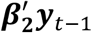 did not suggest any nonstationarity.

Results from the Portmanteau test for serially correlation suggested the presence of serially correlated errors (*p*-value = 0.0035). Inspection of all nine acf and ccf plots of the residuals for lags between -15-to-15 identified the likely reason. The acf plots for Testing and Health Bot showed mild autocorrelation at lag 7. This can be attributed to a “day of the week” seasonal effect. We address this further in the Discussion. Turning our attention towards the normality of the errors, the univariate Jarque-Bera test for Health Bot suggested a departure from this model assumption (*p*-value = 0.0001). This was attributed to the presence of two mild statistical outliers early on in the time series. Since these values were otherwise practically unremarkable and with no assignable cause, we did not remove them.

Figure 4 shows the VECM fit for Census on August 1, 2020. The red line corresponds to the predictions and forecasts (or “fitted values”) from the model, the black dots are the observations, the blue envelope is the approximate in-sample 95% prediction interval band, and the pink envelope is, in this case, the 14-days-ahead out-of-sample forecast interval cone. Up to August 1, the model fit evidences quite reasonable accuracy and precision. Notice we included the 14 actual Census values that were subsequently observed from August 2-to-August 15. The corresponding MAPE is 6.4%. While there is clearly a large outlying value on August 4, the VECM forecast captures the salient feature of the Census counts; that is, a declining local trend. It is interesting to note the declining trend in Testing and Health Bot in late July (Figure 3).

**Figure 4:**
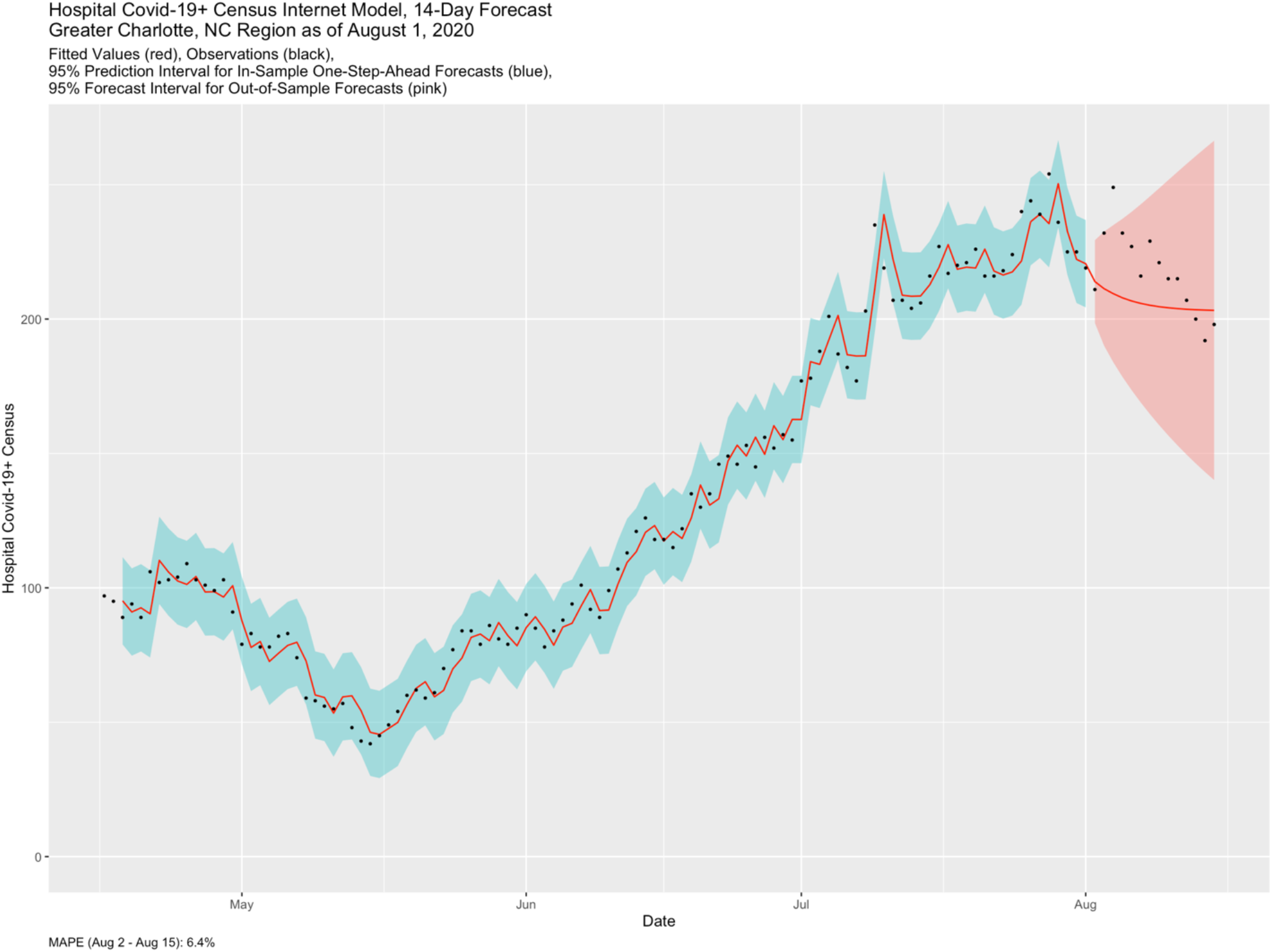
Fitted VECM for Census and 14-Days-Ahead Out-of-Sample Forecast

In Figure 5, we show the distribution of 7-days-ahead out-of-sample MAPE using our time series cross-validation procedure described in the Methods (*n* = 88). The distribution is clearly right-skewed. The median MAPE is 10.5%, while the 95^th^ percentile is 32.9%. In the context of pandemic surveillance and planning, we interpret these results to suggest our MAPE exhibits very good accuracy of the Census forecast, on average. Ceteris paribus, a MAPE beyond 32.9% would be unusual and worthy of further investigation.

**Figure 5:**
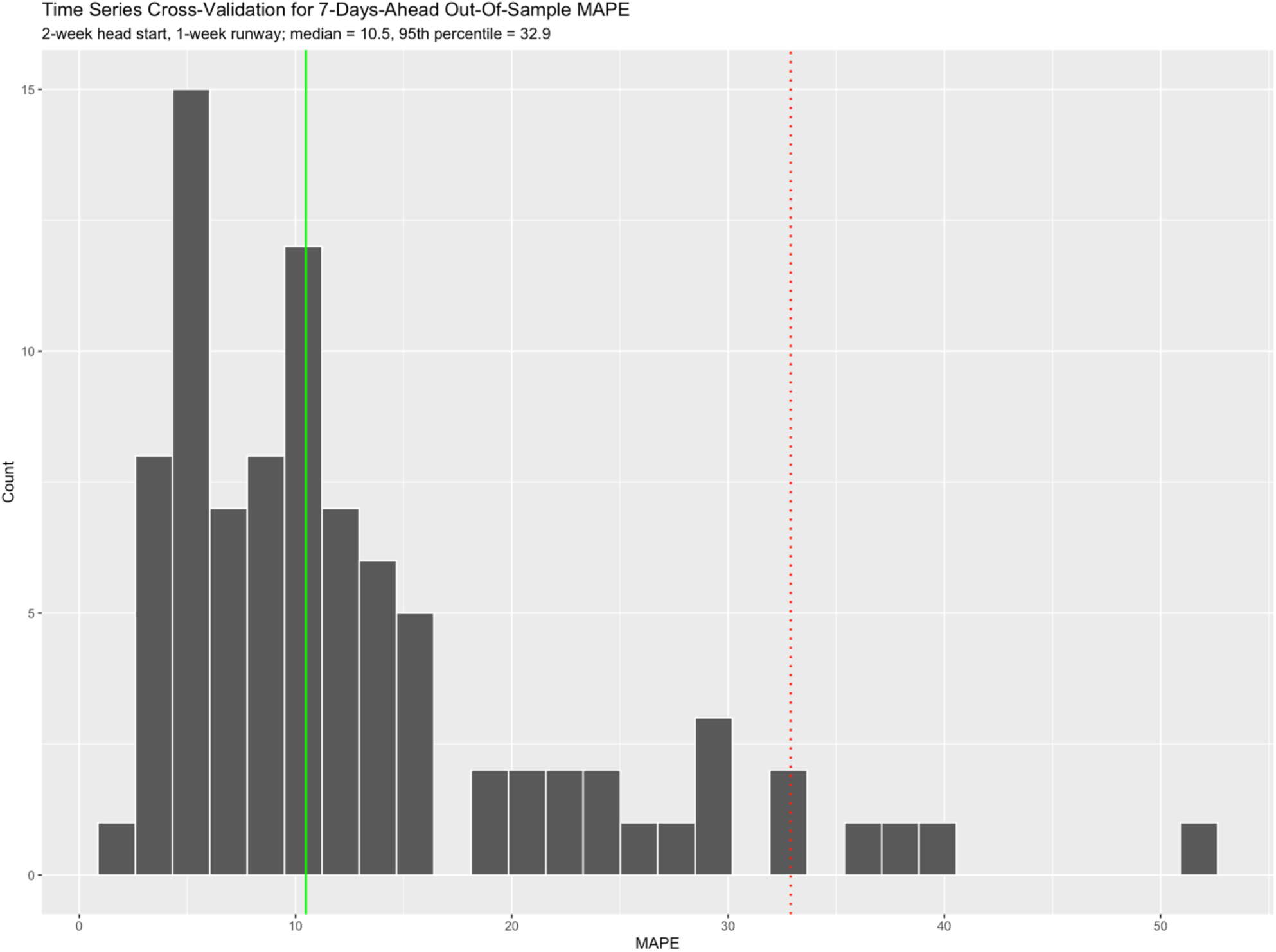
Distribution of 7-Days-Ahead Out-of-Sample MAPE Under Time Series Cross-Validation

When we looked at 7-days-ahead out-of-sample MAPE for the ARIMA model using time series cross-validation, the median MAPE was 8.3%, which was smaller than the value of 10.5% for the VECM. Whether or not this difference is statistically significant would require additional rigor, which is not done here.

## Discussion

In this study, a VECM model inclusive of Internet variables that reflect human behavior during a pandemic performed very well at 7-day forecasting for a regional health system’s COVID-19 hospital census. In terms of short-run fluctuations, there is insufficient evidence that lag 1 values of the three differenced series are useful for prediction of the differenced Census time series. However, in terms of long-run equilibrium, both the error correction terms are statistically significant. Although all three time series, Census, Testing, and HealthBot, are nonstationary, their cointegrating relationships allows to predict the change in Census using the VECM.

There are several advantages in adopting our approach. We have conducted a much more thorough search for candidate Internet variables than what we have observed in the current literature during this pandemic and employed more rigorous statistical methods. Not only have we used Google Trends search terms, but we also evaluated mobility data from Facebook and Apple. Further, we have added data from a healthcare chatbot specifically constructed to assess risk of having COVID-19. Our approach is statistically more rigorous to the extent that we did not stop at stating correlations, but rather provided a formalized multivariate time series model that can be potentially used to provide highly accurate forecasts for health system leaders. Using time series cross-validation in the manner we have described here also provides a way of quantifying forecasting performance for various metrics. The VECM can be easily fit using base R and a few additional packages and we make our code publicly available on GitHub.

The research we have done here can be extended to look at other potential variables that may be leading indicators for predicting COVID-19 Census. These include the community-level effective reproduction number *R*_*t*_ ^54^ and the daily community-level COVID-19 infection incidence, among other examples. Additionally, this same methodology described herein can be extended to look at other health system relevant outcomes, like ICU counts, ventilator counts, or hospital daily admissions. During our specified time period, both the VECM and the ARIMA model provided very good forecasting performance as measured by MAPE, with the VECM returning a slightly larger MAPE value on average. Other performance metrics (e.g., RMSE) were not considered here.

During the 88 days used for this comparison, the Health Bot and Testing time series were relatively stable with respect to linear trend. Using the PELT (Pruned Exact Linear Time) method in the EnvCpt R package, we found two linear trend changepoints for the Health Bot time series (on 05/28/20 and 07/04/20) and one for the Testing time series (on 06/21/20). How the VECM would compare to an ARIMA model if the time series under consideration were to exhibit different types of behaviors would require a further sensitivity study using simulation. We note that just prior to submission (September 26) we refit the VECM and compared it to the ARIMA model. Interestingly, the VECM 7-day forecast projections were trending upward, while those from the ARIMA model were trending downward. A week later, when we computed out-of-sample MAPE for the week of September 27th, we obtained 18.1% for the ARIMA model and 6.8% for the VECM. In fact, Census was beginning to climb. Looking at a multivariate time series plot similar to Figure 3, we observed both Testing and Health Bot start to rise in mid-September and then roughly a week later, Census started to rise.

It is worth mentioning that the VECM is no more or less immune to the same problems we can encounter in obtaining good forecasts when working with any other models. For example, in order to have good forecasts, the future must resemble the past. In the midst of a pandemic, other variables can be introduced with the potential to dramatically alter observed behavior. If a shelter-in-place order, say, were to go in effect in the midst of the forecast horizons and significantly dampen infection spread, then forecasting performance in that time frame would likely suffer. In this scenario, no model will work well.

A potential criticism of our work will likely be that the strong correlations we see between Health Bot and Census, and Testing and Census are “spurious”, being attributable to chance or some underlying unobserved lurking variable. We feel though it is a reasonable assumption that those individuals in the greater Charlotte area that are becoming sick with COVID-19 are likely to search Google for a nearby test site (Testing) or take Atrium Health’s online risk assessment (HealthBot), and then as symptoms subsequently progress proceed to one of Atrium Health’s facilities to be hospitalized.

This study had several limitations. First, in terms of data collection, Google’s designation of the Charlotte NC metro area does not perfectly spatially align with Atrium Health’s core market. Also, Facebook and Apple Map is biased towards users who have enabled their location history on their mobile devices in order to be detected. Second, the time series in this study were not collected using any probabilistic sampling design; rather, they were collected using convenience sampling. Hence, we should be cautious about generalizability of our results. Third, when working with data pulled from the Internet, there is always the chance that the data could be made unavailable or be altered in some way, thus threatening the durability of such models. We were fortunate in that one of our two important Internet variables was from Atrium Health’s own public-facing Microsoft Azure HealthBot, at least in part mitigating this risk for our model. Lastly, perhaps the biggest limitation is that the relationships we have observed in this research could change at any point in the future so that our model is no longer predictive. Stated another way, because these time series are nonstationary, they might not stay in sync over long periods of time as their cross-correlations change.

We initially considered other simpler time series regression models (e.g., autoregressive distributed lag model). However, this approach requires time series under consideration to all be stationary, which ours were not. A spurious regression will result when one nonstationary time series is regressed against one or more other nonstationary time series. Hence, we initially spent a considerable effort trying to stationarize our variables (using differencing, taking logs), and then using lagged versions of the variables before fitting a regression model. In assessing model fit, we were unsuccessful with this approach. Ultimately, the best way to work with nonstationary time series in our case was to acknowledge the cointegration of the variables under study.

Because these are variables derived from the Internet, it would not be unexpected to see evidence of seasonal effects in their time series (e.g., day of the week, weekend versus weekday, etc.). For Testing and Health Bot, we noted the presence of mild autocorrelation in the errors at lag 7. While our VECM results are already very good, incorporating seasonality into our analysis perhaps might improve forecasting performance. What are some options to do this? One approach would be to add seasonal effects directly to the VECM (through ***d***_*t*_). However, with 7 days a week, this would add 18 effect parameters to the model. As we discovered in our case, if many of these effects were unimportant, then this would negatively affect model fit. It is also important to understand that this approach makes the assumption that seasonality is deterministic; whereas, we may actually have stochastic seasonality. A second approach would be to deseasonalize the time series before modeling, i.e., a two-stage approach. We deseasonalized the three time series using seasonal decomposition by loess^55^, noting that the seasonal effects were relatively small. After repeating our data analysis, we found that the VECM fit was not as good. A third approach we leave as a future research topic would be to look at initially fitting a VAR(7) model but disregarding some of the lags (e.g., keeping lags 1 and 7 to address the seasonality, but without the lags 2 to 6, say). This would require more intensive programming in R. With any of these approaches, one still has to check the model for goodness-of-fit and assumptions on the errors; specifically, multivariate normality and lack of serial correlation.

Our VECM model provides a useful forecasting tool that can guide data-driven decision making as health system leaders continue to navigate the COVID-19 pandemic. In exploring candidate predictors, valuable insight was gained as to the relationship between the Internet variables and the hospital census. Both the Health Bot and the Testing time series from the previous 14 days are strongly informative regarding the hospital COVID-19 census and twice gave ample lead time to a substantial change in the census. The VECM provides another model for the hospital COVID-19 positive census in case the simpler ARIMA model no longer exhibits a good fit. It also provides another candidate model that can be used for model-averaged forecasting. While the statistical underpinning of the VECM is somewhat complex, we found the model outputs to be intuitive and thus easily communicated to clinical leaders. Access to this information can help better inform manpower planning and resource allocation throughout the health care system by leveraging insights derived using both of these Internet variables. For these reasons, it is worth considering adding a VECM to the repertoire of a COVD-19 pandemic surveillance program.

## Data Availability

The data and code used in the data analysis is publicly available at GitHub.

https://github.com/philturk/CovCenVECM

## References

1. World Health Organization. IHR Emergency Committee on Novel Coronavirus (2019-nCoV). https://www.who.int/dg/speeches/detail/who-director-general-s-statement-on-ihr-emergency-committee-on-novel-coronavirus-(2019-ncov) (2020).

2. COVID-19 Map. Johns Hopkins Coronavirus Resource Center https://coronavirus.jhu.edu/map.html.

3. Centers for Disease Control and Prevention. Coronavirus Disease 2019 (COVID-19). Centers for Disease Control and Prevention https://www.cdc.gov/coronavirus/2019-ncov/index.html (2020).

4. Jin, Y.-H. et al. A rapid advice guideline for the diagnosis and treatment of 2019 novel coronavirus (2019-nCoV) infected pneumonia (standard version). Mil. Med. Res. 7, 4 (2020).

5. Fowler, J. H., Hill, S. J., Obradovich, N. & Levin, R. The Effect of Stay-at-Home Orders on COVID-19 Cases and Fatalities in the United States. http://medrxiv.org/lookup/doi/10.1101/2020.04.13.20063628 (2020) doi:10.1101/2020.04.13.20063628.

6. Matrajt, L. & Leung, T. Evaluating the Effectiveness of Social Distancing Interventions to Delay or Flatten the Epidemic Curve of Coronavirus Disease. Emerg. Infect. Dis. 26, (2020).

7. Opening Up America Again. The White House https://www.whitehouse.gov/openingamerica/.

8. Public Health Guidance for Reopening. https://www.alabamapublichealth.gov/covid19/guidance.html.

9. Yamana, T., Pei, S., Kandula, S. & Shaman, J. Projection of COVID-19 Cases and Deaths in the US as Individual States Re-open May 4,2020. http://medrxiv.org/lookup/doi/10.1101/2020.05.04.20090670 (2020) doi:10.1101/2020.05.04.20090670.

10. Chopra, V., Toner, E., Waldhorn, R. & Washer, L. How Should U.S. Hospitals Prepare for Coronavirus Disease 2019 (COVID-19)? Ann. Intern. Med. 172, 621–622 (2020).

11. Murthy, S., Gomersall, C. D. & Fowler, R. A. Care for Critically Ill Patients With COVID-19. JAMA 323, 1499–1500 (2020).

12. Ng, K. et al. COVID-19 and the Risk to Health Care Workers: A Case Report. Ann. Intern. Med. 172, 766–767 (2020).

13. Althouse, B. M., Ng, Y. Y. & Cummings, D. A. T. Prediction of Dengue Incidence Using Search Query Surveillance. PLoS Negl. Trop. Dis. 5, e1258 (2011).

14. Husnayain, A., Fuad, A. & Su, E. C.-Y. Applications of Google Search Trends for risk communication in infectious disease management: A case study of the COVID-19 outbreak in Taiwan. Int. J. Infect. Dis. 95, 221–223 (2020).

15. Santillana, M., Nsoesie, E. O., Mekaru, S. R., Scales, D. & Brownstein, J. S. Using Clinicians’ Search Query Data to Monitor Influenza Epidemics. Clin. Infect. Dis. Off. Publ. Infect. Dis. Soc. Am. 59, 1446–1450 (2014).

16. Ocampo, A. J., Chunara, R. & Brownstein, J. S. Using search queries for malaria surveillance, Thailand. Malar. J. 12, 390 (2013).

17. Google Trends. Google Trends https://trends.google.com/trends/?geo=US.

18. Effenberger, M. et al. Association of the COVID-19 pandemic with Internet Search Volumes: A Google TrendsTM Analysis. Int. J. Infect. Dis. 95, 192–197 (2020).

19. Li, C. et al. Retrospective analysis of the possibility of predicting the COVID-19 outbreak from Internet searches and social media data, China, 2020. Euro Surveill. Bull. Eur. Sur Mal. Transm. Eur. Commun. Dis. Bull. 25, (2020).

20. Walker, A., Hopkins, C. & Surda, P. Use of Google Trends to investigate loss-of-smell‒related searches during the COVID-19 outbreak. Int. Forum Allergy Rhinol. 10, 839–847 (2020).

21. Yuan, X. et al. Trends and Prediction in Daily New Cases and Deaths of COVID-19 in the United States: An Internet Search-Interest Based Model. Explor. Res. Hypothesis Med. 5, 1–6 (2020).

22. Nuti, S. V. et al. The Use of Google Trends in Health Care Research: A Systematic Review. PLOS ONE 9, e109583 (2014).

23. Cartenì, A., Di Francesco, L. & Martino, M. How mobility habits influenced the spread of the COVID-19 pandemic: Results from the Italian case study. Sci. Total Environ. 741, 140489 (2020).

24. Jiang, J. & Luo, L. Influence of population mobility on the novel coronavirus disease (COVID-19) epidemic: based on panel data from Hubei, China. Glob. Health Res. Policy 5, 30 (2020).

25. Kraemer, M. U. G. et al. The effect of human mobility and control measures on the COVID-19 epidemic in China. Science 368, 493–497 (2020).

26. Badr, H. S. et al. Association between mobility patterns and COVID-19 transmission in the USA: a mathematical modelling study. Lancet Infect. Dis. S1473309920305533 (2020) doi:10.1016/S1473-3099(20)30553-3.

27. Sasidharan, M., Singh, A., Torbaghan, M. E. & Parlikad, A.K. A vulnerability-based approach to human-mobility reduction for countering COVID-19 transmission in London while considering local air quality. Sci. Total Environ. 741, 140515 (2020).

28. COVID-19 Community Mobility Report. COVID-19 Community Mobility Report https://www.google.com/covid19/mobility?hl=en.

29. Covid-19 Social Distancing Scoreboard — Unacast. https://www.unacast.com/covid19/social-distancing-scoreboard.

30. University of Maryland COVID-19 Impact Analysis Platform. https://data.covid.umd.edu/.

31. COVID-19 - Mobility Trends Reports. Apple https://www.apple.com/covid19/mobility.

32. Facebook Data for Good Mobility Dashboard. COVID-19 Mobility Data Network https://www.covid19mobility.org/dashboards/facebook-data-for-good/ (2020).

33. Bharti, U. et al. Medbot: Conversational Artificial Intelligence Powered Chatbot for Delivering Tele-Health after COVID-19. in 2020 5th International Conference on Communication and Electronics Systems (ICCES) 870–875 (2020). doi:10.1109/ICCES48766.2020.9137944.

34. Ting, D. S. W., Carin, L., Dzau, V. & Wong, T. Y. Digital technology and COVID-19. Nat. Med. 26, 459–461 (2020).

35. Microsoft Health Bot Project - AI At Work For Your Patients. Microsoft Research https://www.microsoft.com/en-us/research/project/health-bot/.

36. Covid19 Symptom Checker. intermountainhealthcare.org https://intermountainhealthcare.org/covid19-coronavirus/covid19-symptom-checker/.

37. WHO Health Alert brings COVID-19 facts to billions via WhatsApp. https://web.archive.org/web/20200323042822/ https://www.who.int/news-room/feature-stories/detail/who-health-alert-brings-covid-19-facts-to-billions-via-whatsapp (2020).

38. Miner, A. S., Laranjo, L. & Kocaballi, A. B. Chatbots in the fight against the COVID-19 pandemic. Npj Digit. Med. 3, 1–4 (2020).

39. Stankiewicz, C. F., Kevin. Apple updated Siri to help people who ask if they have the coronavirus. CNBC https://www.cnbc.com/2020/03/21/apple-updated-siri-to-help-people-who-ask-if-they-have-coronavirus.html (2020).

40. Explore — Opendatasoft. https://demography.osbm.nc.gov/explore/?sort=modified.

41. Google News Initiative Training Center. Google News Initiative Training Center https://newsinitiative.withgoogle.com/training/lesson/6043276230524928?image=trends&tool=Google%20Trends.

42. Centers for Disease Control and Prevention. Coronavirus Disease 2019 (COVID-19). Centers for Disease Control and Prevention https://www.cdc.gov/coronavirus/2019-ncov/index.html (2020).

43. Pfaff, B. Analysis of Integrated and Cointegrated Time Series with R. (Springer-Verlag, 2008). doi:10.1007/978-0-387-75967-8.

44. Dickey, D. A. & Fuller, W. A. Likelihood Ratio Statistics for Autoregressive Time Series with a Unit Root. Econometrica 49, 1057–1072 (1981).

45. Campbell, J. Y. & Perron, P. Pitfalls and Opportunities: What Macroeconomists Should Know about Unit Roots. NBER Macroecon. Annu. 6, 141–201 (1991).

46. Akaike, H. A new look at the statistical model identification. IEEE Trans. Autom. Control 19, 716– 723 (1974).

47. Burnham, K. P. & Anderson, D. R. Model selection and multimodel inference: a practical information-theoretic approach. (Springer, 2002).

48. Johansen, S. Estimation and Hypothesis Testing of Cointegration Vectors in Gaussian Vector Autoregressive Models. Econometrica 59, 1551–1580 (1991).

49. Johansen, S. Likelihood-Based Inference in Cointegrated Vector Autoregressive Models Oxford University Press. N. Y. (1995).

50. Johansen, S. & Juselius, K. Maximum likelihood estimation and inference on cointegration—with appucations to the demand for money. Oxf. Bull. Econ. Stat. 52, 169–210 (1990).

51. Hamilton, J. Time Series Analysis. (Princeton: Princeton University Press, 1994).

52. Zivot, E. & Wang, J. Modeling Financial Time Series with S-Plus®. (Springer New York, 2003). doi:10.1007/978-0-387-21763-5.

53. Hyndman, R. J. & Khandakar, Y. Automatic time series forecasting: the forecast package for R. J. Stat. Softw. 27, (2008).

54. Cori, A., Ferguson, N. M., Fraser, C. & Cauchemez, S. A new framework and software to estimate time-varying reproduction numbers during epidemics. Am. J. Epidemiol. 178, 1505–1512 (2013).

55. Cleveland, R. B., Cleveland, W. S., McRae, J. E. & Terpenning, I. STL: A Seasonal-Trend Decomposition Procedure Based on Loess. J. Off. Stat. 6, 3–73 (1990).

